# Use of consensus term and definition for delayed cerebral ischaemia after aneurysmal subarachnoid haemorrhage

**DOI:** 10.1101/19007260

**Authors:** Matthew J Rowland, Kyle T.S. Pattinson, Mervyn D.I. Vergouwen, Peter J Watkinson

**Affiliations:** Nuffield Division of Anaesthetics, Nuffield Department of Clinical Neurosciences, University of Oxford, John Radcliffe Hospital, Oxford OX3 9DU, United Kingdom; Kadoorie Centre for Critical Care Research, University of Oxford, John Radcliffe Hospital, Oxford OX3 9DU, United Kingdom; Brain Center Rudolf Magnus, Department of Neurology and Neurosurgery, University Medical Center Utrecht, Utrecht University, Utrecht, the Netherlands

## Abstract

**Background and purpose:** In 2010, a multidisciplinary research group proposed a consensus term and definition for the complication of delayed cerebral ischaemia (DCI) following aneurysmal subarachnoid haemorrhage (SAH). We assessed the use of this term and its definition as an endpoint in observational studies and clinical trials.

**Methods:** Firstly, we performed a MEDLINE abstract search from Jan 2008 to Dec 2017 for observational cohort studies and clinical trials to investigate the used terminology over the years. Next, we studied trends in citations of the original paper citing the consensus definitions since publication in 2010.

**Results:** The number of publications citing the 2010 consensus definitions has steadily increased from 18 in 2011 to 54 in 2017. Between 2010 and 2017, 527 papers were published with delayed cerebral ischemia, or another term to describe the same complication, as an endpoint. However, the term delayed cerebral ischemia was used only in 131/527 (25%) of papers and only 14/81 (17%) of clinical trials/cohort studies published in 2017 cited the consensus definitions when outlining study endpoints.

**Conclusions:** Despite publication of consensus terminology and definitions for DCI in 2010, the majority of cohort studies and clinical trials in patients with SAH are not using these. Researchers and editors should be reminded of the importance of using these consensus terminology and definitions in future studies, which will promote the comparability of results between studies, understand the true impact of an intervention, aggregate results in meta-analyses, and construct guidelines with a high level of evidence.

## Introduction

Delayed cerebral ischaemia (DCI) is a complication that occurs in approximately 30% of patients in the first two weeks after aneurysmal subarachnoid haemorrhage (SAH).^1^ Although potentially reversible, it can progress to cerebral infarction and remains the single most important cause of mortality and morbidity in those patients who survive to definitive aneurysm treatment.^2^

Research into DCI has been complicated by inconsistency in the terms used to describe the phenomenon.^3^ These include “cerebral vasospasm”, “delayed ischaemic neurological deficit”, “secondary ischaemia” and “symptomatic vasospasm” amongst many others. The major cause of this confusion arises from the combining of radiological evidence of vasoconstriction after SAH (so called “vasospasm”) with the clinical features of neurological deterioration due to delayed cerebral ischaemia (DCI). Although SAH is associated with cerebral vasoconstriction^4,5^ and cerebral infarction after SAH is strongly associated with poor clinical outcomes,^6,7^ the exact contribution of cerebral vasoconstriction in DCI remains unclear as the two often do not co-exist.^8^

In 2010 a consensus statement was published in Stroke from a multidisciplinary group of research experts which proposed a standardised term and definitions for clinical deterioration and cerebral infarction due to DCI, to be used as an endpoint in future observational studies and clinical trials of SAH (see Supplementary Material).^9^ The importance of these standardised term and definitions was further emphasised through the publication of a consensus statement on the critical care management of patients with SAH in 2011.^10^ We sought to quantify the effect of the publication of this standardised term and its definitions in clinical studies of SAH.

## Methods

First, an electronic literature search was performed using the Medline database from January 2008 to December 2017 to investigate the used terminology in cohort studies and clinical trials over the years. The search criteria covered four terms: “subarachnoid haemorrhage”, “delayed cerebral ischaemia”, “vasospasm”, “cerebral infarction” and “delayed ischaemic neurological deficit” with appropriate synonyms and spelling variations. To ensure complete coverage, both MeSH and free text terms were used (see Supplementary Material for full search term strategy). Studies included were observational, cohort or clinical trials in patients with aneurysmal SAH both prospective and retrospective studies. All publications were limited to those involving human subjects. Exclusion criteria were: 1) duplicates; 2) manuscripts not in the English language; 3) abstracts, case reports, conference presentations, editorials, meta-analyses, reviews, and expert opinions; and 4) papers specifically dealing with definitions of vasospasm, DCI etc. Identified papers were screened by title and abstract for suitability by one author (MR).

Following this, the full text of each paper selected for inclusion was reviewed. Papers were included if DCI, or another term to describe this complication, was used as an endpoint in the study. The following data were recorded from included papers:

1. Study date range (if documented)
2. Endpoint(s) terminology (e.g. DCI, angiographic vasospasm, DIND etc.)
3. Whether the consensus definition for clinical deterioration or cerebral infarction due to DCI (Vergouwen et al. 2010 or via Diringer et al. 2011) was referenced?
4. If not, whether any alternative reference was used for the defined endpoint(s) used in the study.

Finally, SCOPUS Web of Science was searched to quantify the number of citations of the original consensus definitions proposed by Vergouwen et al.^9^ between publication in 2010 and December 2017.

## Results

**Figure 1** shows the total number of papers retrieved from Scopus Web of Science citing the consensus definitions from Vergouwen et al.^9^ from January 2010 to December 2017. Over that period, there was an increasing recognition of the 2010 consensus definitions with a total of 326 citations from 289 publications over that period, steadily increasing over time.

**Figure 1:**
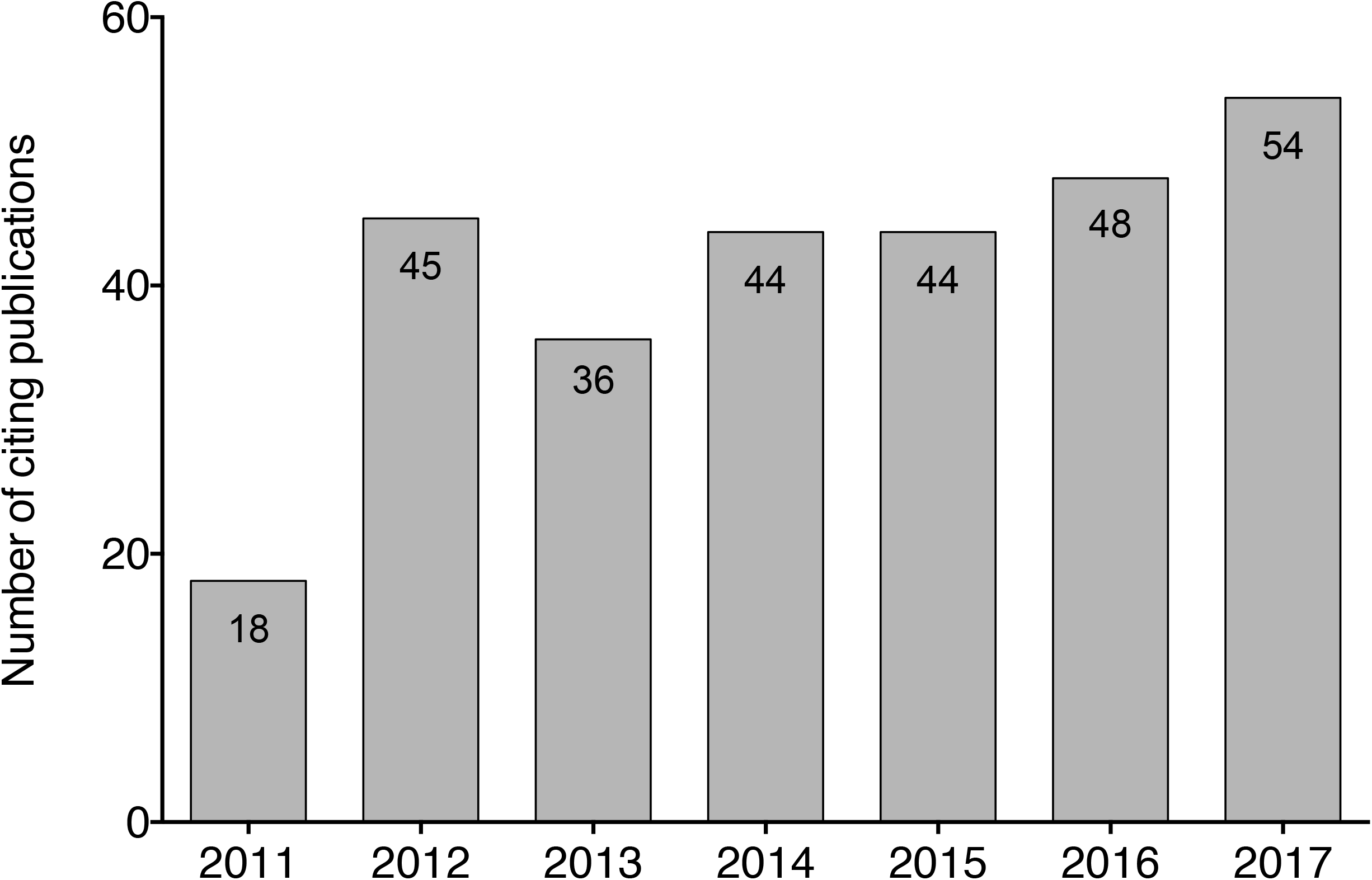
PRISMA flowsheet for the literature search to identify studies for inclusion in the analysis

**Figure 2A** shows the complete adapted PRISMA flow diagram for inclusion of papers into the analysis. 3130 papers were retrieved from the initial literature search. From this, 449 full text papers were identified for inclusion in the final search as per the criteria above. Further to the original search criteria and during final analysis, three papers were subsequently excluded as they used data from multiple different trials over a long time-period using different definitions of DCI (PMID 27717776, 28862545 and 27717776). Finally, the full text for one paper was not possible to find on the publisher’s website or via web searches (PMID 21156426).

**Figure 2:**
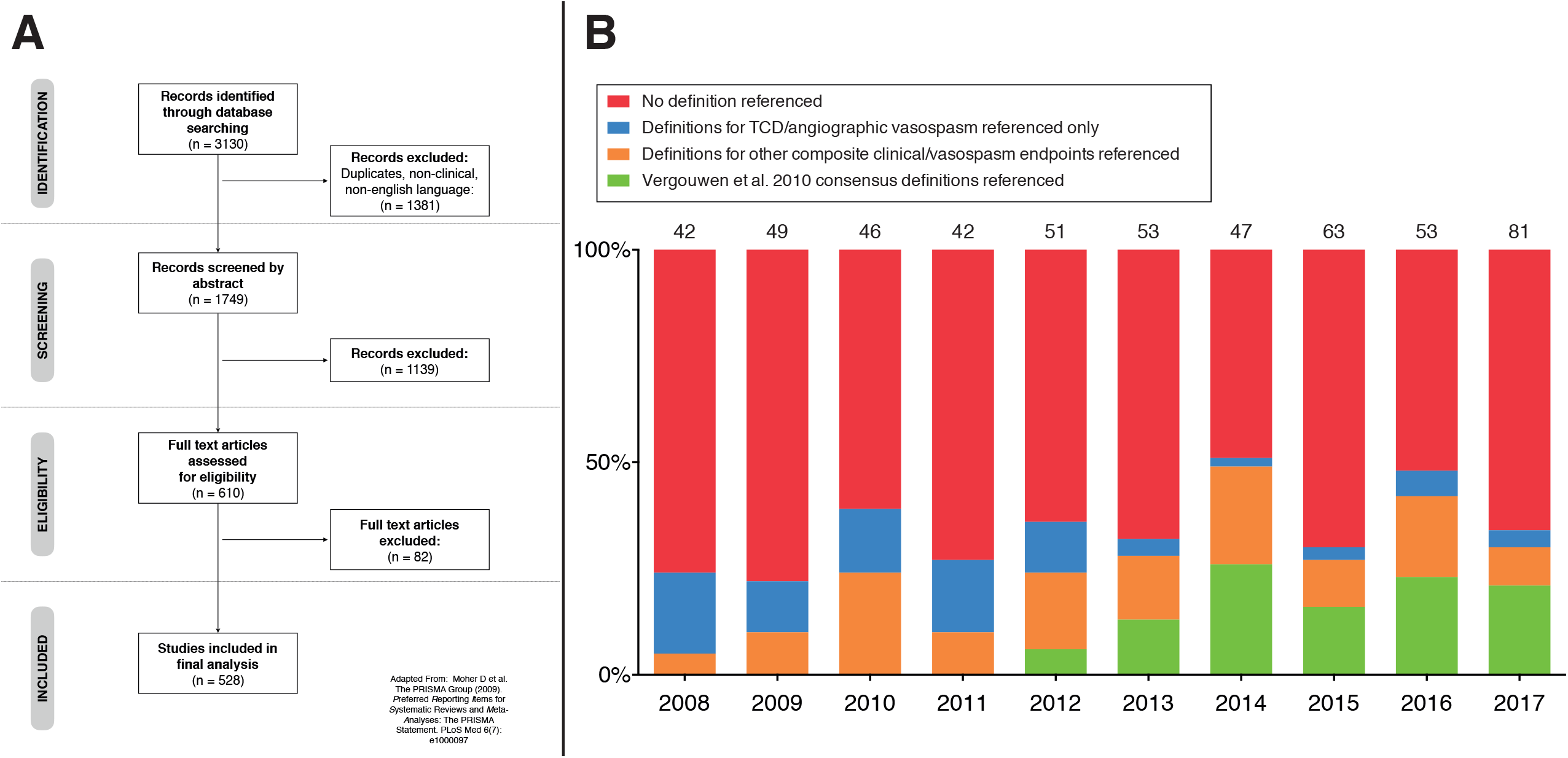
**A** – Number of publications citing the Vergouwen et al. 2010 consensus definitions by year from 2011 – 2017. **B** – Percentage of all publications between 2008 and 2017 citing either the 2010 consensus definitions, definitions for trans-cranial Doppler (TCD) endpoints, any other composite clinical/radiological definition or not citing defined end points at all

**Figure 2B** shows the proportion of retrieved papers defining study endpoint using the consensus definitions from 2010, a reference for trans-cranial Doppler only, any other reference for composite clinical/radiological endpoints, or no reference at all by year of publication. Although the number of publications citing the 2010 consensus definitions has steadily increased from 18 in 2011 to 54 in 2017, the term “delayed cerebral ischemia” was only used in 131/527 (25%) of papers. Furthermore, only 14/81 (17%) of clinical trials/cohort studies published in 2017 cited the consensus definitions when outlining study endpoint with 23/81 (28%) of publications published in this period not citing any definition used as an endpoint.

Finally, **Table 1** shows the total number of papers by year included in the final analysis, sub-divided by the type of papers published (e.g. clinical trial, prospective cohort study etc.) and by the range of different terms used to describe the clinical endpoint being studied. In 2017, only 39/81 (48%) of cohort studies/clinical trials still used the term “vasopasm” as an endpoint – either defined with transcranial Doppler or angiography, clinical characteristics (such as “symptomatic” or “clinical”) or not defined in the manuscript. Seven papers cited the consensus definitions by Vergouwen et al. but either used incorrect terminology in describing the end-point being cited (such as “delayed ischaemic neurological deficit (DIND)”, “clinical vasospasm” and “symptomatic vasospasm”) or linked “angiographic vasospasm” to the cited definition for DCI. A number of papers also incorrectly used the abbreviation DCI to mean “delayed cerebral infarction” (e.g. PMID: 27518526, 28946178 and 28548598)

**Table 1:** Number of papers by type and by defined study endpoint(s) from 2008 – 2010

|  | 2008 | 2009 | 2010 | 2011 | 2012 | 2013 | 2014 | 2015 | 2016 | 2017 |
| --- | --- | --- | --- | --- | --- | --- | --- | --- | --- | --- |
| **Total number of papers** | 42 | 49 | 46 | 42 | 51 | 53 | 47 | 63 | 53 | 81 |
| Clinical trials | 4 | 5 | 3 | 4 | 5 | 5 | 4 | 5 | 3 | 4 |
| Prospective cohort studies | 31 | 28 | 27 | 23 | 34 | 34 | 26 | 44 | 27 | 46 |
| Retrospective cohort studies or analysis of RCT | 7 | 16 | 16 | 15 | 12 | 14 | 17 | 14 | 23 | 31 |
| **Study Endpoints** |  |  |  |  |  |  |  |  |  |  |
| Consensus term and definition | 0 | 0 | 0 | 0 | 3 | 7 | 12 | 10 | 12 | 9 |
| Delayed cerebral ischaemia (DCI) (non-consensus definition) | 2 | 6 | 4 | 7 | 10 | 7 | 6 | 14 | 14 | 26 |
| Cerebral infarction or variant (non-consensus definition) | 2 | 8 | 6 | 5 | 3 | 4 | 1 | 2 | 5 | 10 |
| Vasospasm (total) | 52 | 50 | 41 | 25 | 38 | 39 | 36 | 36 | 30 | 39 |
| Vasospasm alone | 1 | 0 | 2 | 5 | 9 | 3 | 6 | 6 | 11 | 9 |
| Symptomatic vasospasm | 7 | 11 | 13 | 9 | 8 | 10 | 7 | 7 | 8 | 11 |
| Clinical vasospasm | 7 | 5 | 2 | 2 | 1 | 2 | 1 | 7 | 1 | 4 |
| TCD vasospasm | 20 | 18 | 17 | 5 | 12 | 7 | 7 | 8 | 4 | 5 |
| Angiographic vasospasm | 17 | 16 | 7 | 4 | 8 | 9 | 8 | 8 | 6 | 10 |
| Delayed ischaemic neurological deficit (DIND or variant) | 9 | 9 | 4 | 3 | 6 | 8 | 4 | 4 | 3 | 8 |

## Discussion

The results from this study demonstrate that despite the publication of standardised consensus definitions for both clinical deterioration and cerebral infarction due to DCI in 2010 by a multidisciplinary group of expert researchers, there has been poor uptake and citation of these definitions in subsequent published studies in patients with SAH. Although the number of studies using DCI as a defined endpoint has increased, there still remains a wide number of alternative terms used to describe both the clinical and radiological features investigated in studies. This continues to hamper interpretation of research into SAH due to the ongoing confusion between radiological evidence of vessel narrowing (“vasospasm”) and clinical neurological deterioration due to DCI.

The recommendation from the 2010 consensus paper was that the proposed definitions “be used in future clinical trials and observational studies that have DCI as an outcome event until more accurate, reliable and sensitive tests are developed to measure DCI”.^9^ Whilst a lag in the inclusion of these definitions is to be expected – especially in the case of trials commenced prior to 2010 – the proportion of papers citing them has remained between 6 and 26% which is disappointing and serves to make meta-analysis or systematic review of these studies/trials extremely difficult.

Another important aspect of the consensus definitions was that the authors stressed the occurrence of clinical deterioration due to DCI be described separately from radiological or TCD evidence of cerebral vasoconstriction. As can be seen from Table 2, there remains a multitude of terms used to define study endpoints investigating secondary clinical deterioration following SAH. These are often composite endpoints including both clinical features of neurological deterioration (variously defined and often un-referenced) as well as direct or indirect measures of cerebral vasoconstriction such as cerebral angiography or trans-cranial Doppler ultrasound. Furthermore, a number of papers retrieved from this search were noted to use clinical deterioration due to DCI as an endpoint and cite the consensus definitions but confuse the definition of DCI with “vasospasm”. Examples of this include “DCI was defined as a symptomatic vasospasm, infarction attributable to vasospasm, or both (Frontera et al 2009, Vergouwen et al 2010)” and “Symptomatic vasospasm was defined as neurological deterioration with documented arterial vasospasm (Frontera et al 2009, Vergouwen et al 2010).” Finally, a number of papers cited a definition similar to the consensus definitions but subtly different e.g. “Delayed cerebral ischemia (DCI) was defined as (1) clinical deterioration (i.e. a new focal deficit, decrease in level of consciousness, or both), and/or (2) a new infarct on CT that was not visible on the admission or immediate postoperative scan, when the cause was thought by the research team to be vasospasm. Other potential causes of clinical deterioration or CT lucencies, such as hydrocephalus, rebleeding, cerebral edema, retraction injury, ventriculitis, metabolic derangements, and seizures were rigorously excluded”. In this example, there is no clarification of what level of decreased consciousness to consider nor duration.

The factors underlying the poor uptake of these consensus definitions remain difficult to isolate. They were produced by a multi-disciplinary group of clinical and research experts and published open access in a high impact journal so should be widely available for reference. A number of studies published since 2010 – especially clinical trials - will have developed protocols and commenced recruitment prior to the publication date and therefore would not be expected to have used the consensus definitions. Furthermore, a number of papers will have cited the paper by JA Frontera et al. from 2009 which was one of the first to address what is the most clinically relevant definition for vasospasm and highlighted the importance of using the term DCI. However, the definition used for DCI in that paper is slightly different to the one used in the consensus definitions that were subsequently published in 2010. It is essential that those editing papers for publication and reviewing grants reinforce the importance of careful definition of study endpoints to allow researchers to both interpret data clinically and to allow inclusion of studies in future meta-analysis/systematic review.

## Conclusion

This study has highlighted the ongoing heterogeneity in terminology and definition of DCI. Consistent use of the clear consensus definitions for both clinical deterioration and cerebral infarction due to DCI is essential in order to collect and interpret useful data from clinical studies of patients with SAH. Authors need to be reminded that neurological deterioration due to DCI and cerebral infarction should be separated from radiological vasoconstriction when used as an endpoint for SAH research. The term “vasospasm” should be restricted to describing radiological abnormalities in blood vessels seen after SAH. Cerebral infarction remains the preferred endpoint for use in clinical trials investigating DCI after subarachnoid haemorrhage.

## Data Availability

not applicable

## Supplementary Material

### 1. Definitions

Proposed definitions of DCI and cerebral infarction after SAH for use as an outcome event in clinical trials and observational studies (Vergouwen et al. 2010)

**Clinical deterioration caused by DCI**

The occurrence of focal neurological impairment (such as hemiparesis, aphasia, apraxia, or neglect), or a decrease of at least 2 points on the Glasgow coma scale (either on the total score or on one of the components)

This should

1. Last for at least 1 h
2. Not be apparent immediately after aneurysm occlusion
3. Not be attributed to other causes by means of clinical assessment, CT or MRI scanning of the brain, and appropriate laboratory studies

**Cerebral infarction**

The presence of cerebral infarction on either:

1. CT or MRI scan of the brain within 6 weeks after SAH
2. The latest CT or MRI scan made before death within 6 weeks after SAH
3. Proven at autopsy

This evidence must not be present on the CT or MRI scan between 24 and 48 h after early aneurysm occlusion and must not be attributable to other causes such as surgical clipping or endovascular treatment

Hypodensities on CT imaging resulting from ventricular catheter or intraparenchymal haematoma should not be regarded as cerebral infarctions from DCI

### 2. Search term strategy

SUBARACHNOID HEMORRHAGE

(subarachnoid and haemorrhag*).af.

(subarachnoid and hemorrhag*).af.

(SAH or aSAH).af.

CEREBRAL INFARCTION

VASOSPASM

delayed cerebral”.af.

“delayed ischaem*”.af.

“delayed ischem*”.af. (DCI or DIND).af.

“vasospasm*”.af.

(DIND)

“delayed ischaemic neurological deficit”.af

“delayed ischemic neurological deficit”.af

